# Development of an Optical Assay to Detect SARS-CoV-2 Spike Protein Binding Interactions with ACE2 and Disruption of these Interactions Using Electric Current

**DOI:** 10.1101/2020.11.24.20237628

**Authors:** Mahmoud Al Ahmad, Farah Mustafa, Neena Panicker, Tahir A. Rizvi

## Abstract

This study proposes a novel optical method of detecting and reducing SARS-CoV-2 transmission, the virus responsible for the COVID-19 pandemic that is sweeping the world today. SARS-CoV-2 belongs to the β-coronaviruses characterized by the crown-shaped spike protein that protrudes out of the virus particles, giving the virus a “corona” shape; hence the name coronavirus. This virus is similar to the viruses that caused SARS (severe acute respiratory syndrome) and MERS (Middle East respiratory syndrome), the other two coronavirus epidemics that were recently contained within the last ten years. The technique being proposed uses a light source from a smart phone and a mobile spectrophotometer to enable detection of viral proteins in solution or paper as well as protein-protein interactions. The proof-of-concept is shown by detecting soluble preparations of spike protein subunits from SARS-CoV-2, followed by detection of the actual binding potential of the spike protein with its host receptor, the angiotensin-converting enzyme 2 (ACE2). The results are validated by showing that this method can detect antigen-antibody binding using two independent viral protein-antibody pairs. The binding could be detected optically both in solution and on a solid support such as nitrocellulose membrane. Finally, this technique is combined with DC bias to show that introduction of a current into the system can be used to disrupt the antigen-antibody reaction, suggesting that the proposed extended technique can be a potential means of not only detecting the virus, but also reducing virus transmission by disrupting virus-receptor interactions electrically.

**Significance:** The measured intensity of light can reveal information about different cellular parameters under study. When light passes through a bio-composition, the intensity is associated with its content. The nuclei size, cell shape and the refractive index variation of cells contributes to light intensity. In this work, an optical label-free real time detection method incorporating the smartphone light source and a portable mini spectrometer for SARS-CoV-2 detection was developed based on the ability of its spike protein to interact with the ACE2 receptor. The light interactions with control and viral protein solutions were capable of providing a quick decision regarding whether the sample under test was positive or negative, thus enabling SARS-CoV-2 detection in a rapid manner.

## Introduction

The world is currently facing the COVID-19 pandemic caused by the appearance of a novel coronavirus in the human population towards the end of 2019^1^. Within only a few months, this virus had spread to most countries of the world, infecting millions (> 21 million as of Aug. 17 2020) and causing > 770,000 deaths^2^. Unfortunately, the complete clinical picture of COVID-19 is not yet fully known and most likely depends upon a number of factors, including both the virus as well as the host characteristics. Successful detection of SARS coronavirus 2 (SARS-CoV-2) plays an important role in stopping its spread. At the moment, oropharyngeal and nasopharyngeal swabs are primarily used for virus detection. However, it is not clear how many virus particles of SARS-CoV-2 are needed to trigger an infection. It is also not clear whether the virus load in an infected individual is correlative of disease severity since both asymptomatic individuals and patients with COVID-19 symptoms can demonstrate similar viral loads^3^. It has been estimated that SARS-CoV-2 exhibits a higher rate of virus replication compared to SARS-CoV-1, which can increase disease transmissibility^4^. Statistically, confirmed COVID-19 cases worldwide are 100 times higher than the confirmed cases of SARS and MERS^5^. This is because, 1) SARS-CoV-2 replicates at much higher levels in the nose and mouth than SARS and MERS, and 2) this leads to much higher levels of virus shedding in the environment by people who are either pre-symptomatic or asymptomatic. Thus, a large percentage of infected people can transmit the virus without realizing that they are even infected^6^. Due to these reasons, fast, cheap and accurate methods of SARS-CoV-2 detection is the need of the hour and should slow the spread of the virus till a vaccine or effective therapy can be found.

Rapid detection methods independent of lab setting have been identified as one of the foremost priorities for promoting epidemic prevention and control. Currently, the molecular technique of quantitative real time polymerase chain reaction (qRT PCR) is the gold standard for SARS-CoV-2 detection using samples from respiratory secretions^7^. However, it is a time consuming and cumbersome procedure that takes long processing times over days for results^8^. Several other molecular assays have been developed to detect SARS-CoV-2, such as enzyme-based assays like ELISAs, and rapid tests that aim to detect either antibodies against the virus or the viral antigen themselves^4^. Nevertheless, most of these antigen-antibody-based assays have failed quality control due to their rapid development without proper testing and result in either false negative or false positive detection due to the long time it takes to develop serum responses to the viral infection (from days to weeks)^9^. Thus, most of the methods used so far either require skilled manpower and are time consuming if accurate, or not reliable at all, if fast. On the other hand, biosensor technology provides excellent sensitivity, but have their own caveats. For example, some biosensors require metal coating deposited on the device, thereby raising cost^10^, while other suffer from temperature-dependence which can be a hindrance for portable biosensors in outdoor conditions^11^. Some require expensive reagents and reaction times that are often longer^12^. Mavrikou et al. have used Bioelectric Recognition Assays along with artificially-engineered cells to demonstrate direct detection of SARS-CoV-2 surface antigens without prior sample processing^13^; however, they still have to show whether their system will work in the real-world scenario. Optical, label-free biosensors have been utilized frequently in biomolecular detection due to their ability for continuous monitoring and high sensitivity to local variation, including the refractive index change^14^. They are capable of detecting interactions between molecules and their surrounding media^15^.

In terms of detection, the most prominent feature of the SARS-CoV-2 virus, like other coronaviruses, is the spike protein (S) that protrudes out of the virus particle essentially like “spikes” as the name suggests The spike protein forms a trimer that is used by the virus to enter susceptible cells using the angiotensin-converting enzyme 2 (ACE2) protein as the cellular receptor^16^, the same protein used by the SARS-CoV-1 virus that caused the first SARS epidemic in 2003 (Fig. 1(a)). The spike protein is cleaved by host proteases into two subunits: the surface subunit S1, and the transmembrane subunit S2^17^ (Fig. 1(b)). It is the surface S1 subunit that is used by the virus to interact with ACE2 protein using its receptor binding domain (RBD)^18^. This allows the virus to attach to the susceptible cells, while the S2 protein is used for the actual fusion of the virus with the cell membrane, allowing the virus to be endocytosed and release its genomic RNA cargo, wrapped up in the nucleocapsid protein (NCP), into the cytoplasm^19,20^. The viral genomic RNA is immediately used to translate viral proteins that are used for successful virus replication in the susceptible cells^21^. The spike protein is also one of the most immunogenic proteins of the virus towards which most of the neutralizing antibody responses against the virus are generated in infected individuals, making it an ideal candidate for vaccine as well as a target of drug development^22,23,24^. In the context of COVID-19, it has been observed that the SARS-CoV-2 spike glycoprotein binds ACE2 with 10-20-fold higher affinity than SARS-CoV-1 spike protein, which may explain the higher transmissibility of SARS-CoV-2 in the human population^25^.

**Figure 1:**
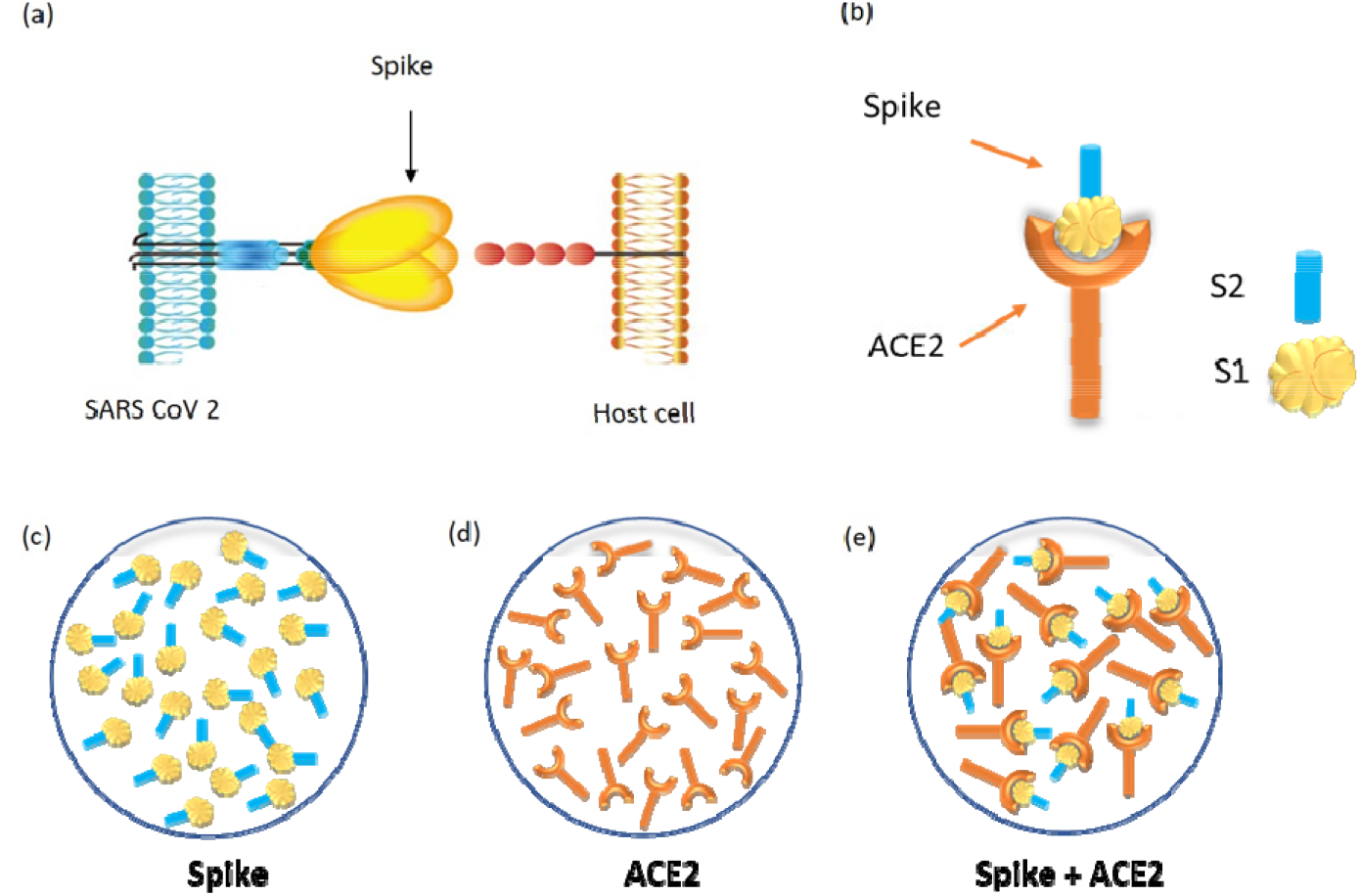
Schematic illustration of SARS-CoV-2 spike protein and ACE2 receptor binding. **(a)** SARS-CoV-2 binding to the ACE2 receptor on the host cell surface. **(b)** Binding of ACE2 and spike protein along with illustration of the spike protein subunits, S1 and S2. **(c)** Schematic showing the distribution of the spike protein in solution. **(d)** Schematic showing the distribution of ACE2 in suspension. **(e)** Distribution of ACE2 and S protein after binding in solution.

The receptor binding domain of SARS-CoV-2 is the key region of the S protein that affects virus spread. Xia et al. have confirmed these observations by showing that even the fusion capability of SARS-CoV-2 S2 subunit is better than that of SARS-CoV-1, further explaining the increased infectivity of the virus compared with other corona viruses^26^. They further show that lipopeptide inhibitors can be developed that can disrupt such fusion capability to inhibit the ability of the virus to infect cells^27^. Similarly, Seydoux et al. have shown the utility of isolating S-specific antibody-producing B-cell clones from COVID-19 patients^28^. They further demonstrate that the most potent amongst these antibodies was targeted against the RBD of the S protein which was able to block the interaction of the S protein with ACE2 successfully. Yang et al. have tested several binding inhibitor peptides targeting the virus early attachment stages^29^. Others have observed a strong correlation between levels of RBD-binding antibodies and SARS-CoV-2 neutralizing antibodies in patients^22,30^. Thus, study of spike protein interaction with the ACE2 receptor can be of importance for not only virus entry into cells, but also as a means of inhibiting virus infection of susceptible cells, development of vaccines, as well as for detection of virus infection.

In this study, we demonstrate the use of a light-based method to detect SARS-CoV-2 and potentially disrupt its binding ability with its receptor, rendering the virus non-infectious by combining optical detection with electric current. The measured intensity of light can reveal information about different cellular parameters under study. Light characteristics can be correlated with the contents of the sample under test and reflect the complexity of their exterior or interior structures^31^. When light passes through a bio-composition, the intensity is associated with its content^32^. The nuclei size, cell shape and the refractive index variation of cells contributes to light intensity^33^. It is worth adding that the measured optical spectrum consists of many features that can reveal important information about the sample under test^34^. In this work, an optical label-free real time detection method incorporating the smartphone light source and a portable mini spectrometer for SARS-CoV-2 detection was developed based on the ability of its spike protein to interact with the ACE2 receptor. The light interactions with control and viral protein solutions were capable of providing a quick decision regarding whether the sample under test was positive or negative, thus enabling SARS-CoV-2 detection in a rapid manner.

## Results and discussions

### Experimental Design

The experimental setup utilized in this study is shown in Fig. 2(a), incorporating a mini spectrometer and a smart mobile phone that was employed as a light source with its power spectrum depicted in Fig. 2(b). The measured optical power of the beam exhibited maximum power at a wavelength of 623 nm^35^. The mini-spectrometer C11708MA (Hamamatsu/Japan) was used to measure the light intensity as it passes through test substances with spectral response ranging from 640 to 1010 nm^36^. The wavelength reproducibility was between −0.5 to 0.5 nm and a maximum of 20 nm FWHM spectra, under constant light conditions. The sample under test was placed between the mobile light source and the mini-spectrometer, as shown in Fig. 2(a). The measurements were conducted with the room lights on. The distances between the light source, the spectrometer, and the sample holder were adjusted to eliminate any possible interference and to stabilize the spectrometer performance. Furthermore, the spectrometer was aligned with the light source and sample cuvette to achieve a straight path of light. Figure 2(c) illustrates the incident, reflected, and transmitted light intensities. The light intensities were linked through the Kirchhoff’s Law of Radiation^37^, which correlates the optical absorbance, transmittance, and reflection along with the incident wave.

**Figure 2:**
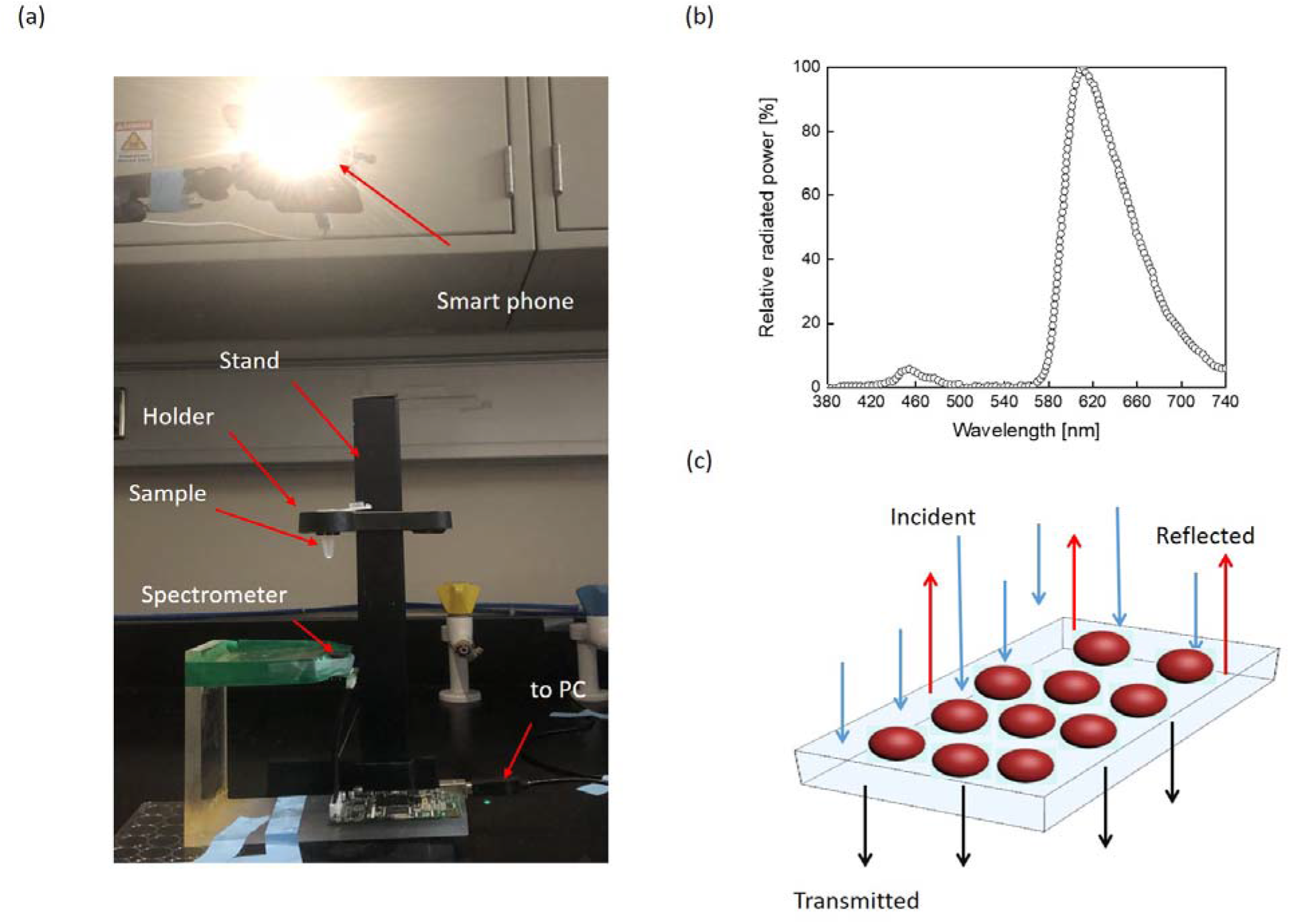
The proposed concept of optical detection and the experimental design: **(a)** The optical measurement setup is shown consisting of a smart phone as a light source and the mini-spectrometer utilized to collect the light waves passing through the sample kept in the holder. **(b)** The smart phone power spectra *versus* wavelength. **(c)** Illustration of the spectrometer detection principle.

This experimental setup was first used to characterize the two spike proteins subunits, S1 and S2 that are encoded by all coronaviruses and, as mentioned, allow virus entry into susceptible cells (Fig. 1(b)). Figure 3(a) shows the optical responses for both proteins along with their corresponding blank samples. The measured optical intensity changed from 600 to 750 nm, within the light source spectrum measured earlier in Fig. 2(b). The response of the blank samples was performed first, followed by the two protein suspensions, the responses to which were recorded individually as shown in Fig. 3(a).

**Figure 3:**
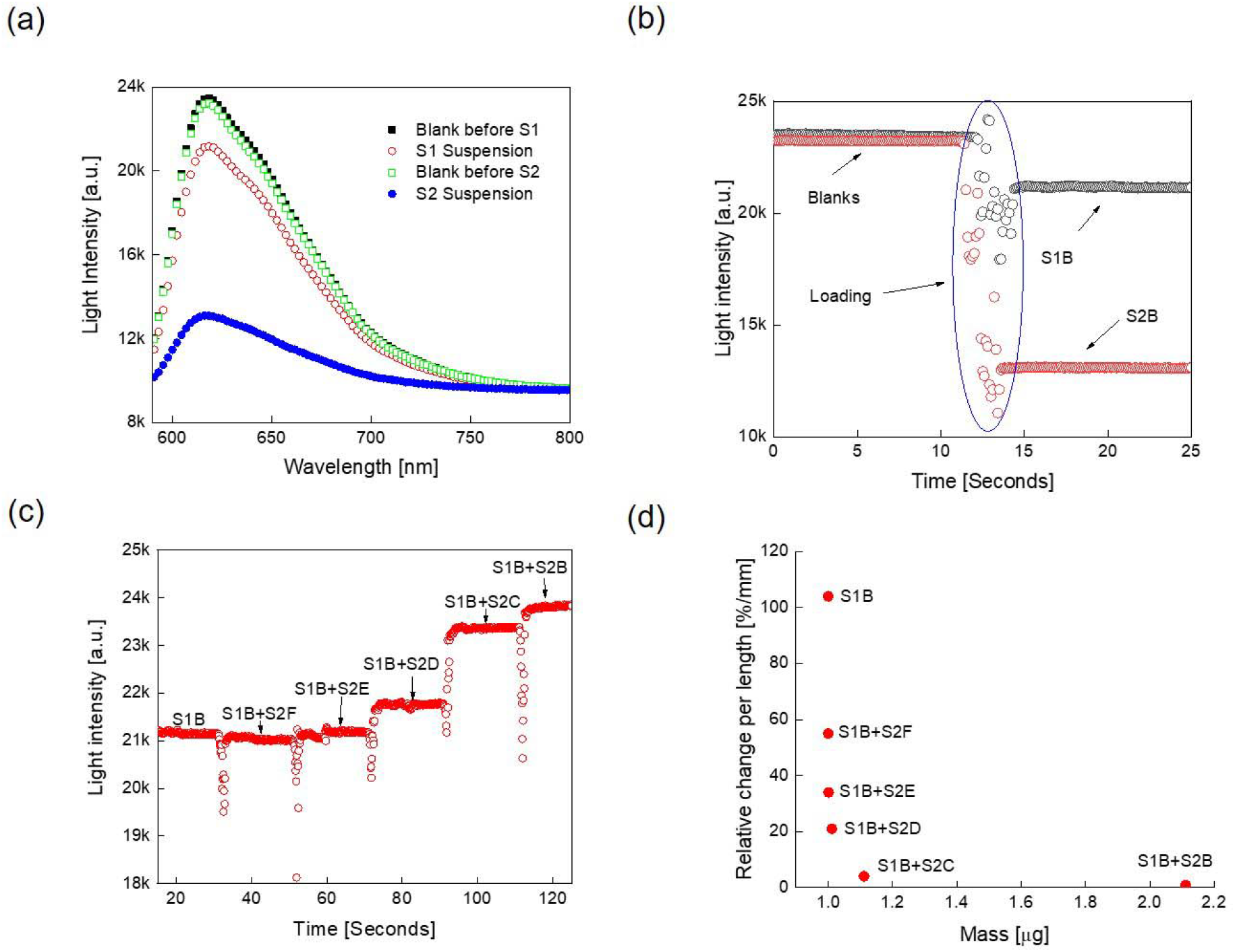
Optical measurements of the spike protein subunits S1 and S2: **(a)** Measured responses for spikes proteins S1 and S2 at the highest concentration individually (S1B and S2B, respectively), along with their corresponding blanks. **(b)** Time domain measurements of the microcentrifuge tube, the blank (shown in gray circles) *versus* water (red circles) at a wavelength of 623 nm. **(c)** Measured optical responses for the mixed protein samples *versus* time. Samples S1B and S2B were at 5000 copies per ml, S2C, S2D, S2E and S2F are the serial dilutions of S2B at 10-, 100-, 1,000- and 10,000-fold, respectively. **(d)** Relative change in light intensity per light path *versus* loaded mass. All optical responses were measured at 623 nm. Light intensity was measured as arbitrary units (a.u).

Figure 3(a) reveals that S2 exhibited a higher “back scattering” and/or absorbance than S1. The response of the two blank samples was quite comparable, showing the reproducibility of the results. Since the maximum difference between the blank and the two protein samples was observed at 623 nm, this wavelength was chosen for further experimentation, which is also the wavelength at which the optical power of the smart phone is at its maximum.

### Optimization of the Sample Reading Conditions

Initial test of this experimental setup revealed that it had one major drawback; i.e., when samples were loaded into the holder, the angle and position of the microcentrifuge tube changed which affected the results obtained. To ensure that the results were reproducible, the measurements for the same samples were conducted over different days and on each day the setup was standardized since the position of the mobile phone, spectrometer and samples could vary. To overcome this caveat and have more consistence measurements without constant standardization, advantage was made of the ability of the spectrometer to provide light intensity measurements over time. Hence, after placing the microcentrifuge tube into the holder, the measurement mode would be started and the corresponding “blank” recorded. Then the sample would be added after ∼ 100 mSecond, while keeping the measurement mode on. Figure 3(b) illustrates the corresponding measurement profile for S1B and S2B individual samples suspended in water over time. Initially, a fluctuation in the light intensity was observed with time as each sample was added to the tube, but then it stabilized with time. As expected, the blank exhibited the maximum measured light intensity, while the suspended samples showed lower light intensity than the blank once stabilized.

### Test of the Spike Proteins Using the Proposed Experimental Set Up

To test the proof-of-principle, initially a mixing experiment was conducted at a light wavelength of 623 nm. Towards this end, 250 μL of S1B protein solution was tested at the same maximum concentration at 5,000 copies/mL followed by addition of same amount of S2B. Figure 3(c) shows the light intensity (as arbitrary units, a.u.) with time as the protein samples were added to the transparent measurement container in a sequential manner. This was followed by addition of 250 μL of ten-fold serial dilutions of the S2 protein at equal time intervals to the S1B + S2B samples. As can be seen from Fig. 3(c), with the addition of the S2 protein, the light intensity increased. The biggest increase was observed with the concentrated S2B sample followed by its ten-fold dilution samples S2C, S2D, S2E, etc., until S2F addition as a 1:10,000 dilution had no extra effect on the increase in light intensity, revealing the limit of detection of the assay (5000 molecules per mL x 250 µl x 1/10,000 = 125 molecule per mL). These results reveal that the ratio between the S1 and S2 protein concentration plays an important role in the light intensity levels measured. The ratio of S1 and S2 in the virus is the same since both originate from the cleavage of S protein. However, the S1 subunit is expressed on the cell surface, while the S2 subunit is embedded in the lipid bilayer of the cell membrane; therefore, S2 is less available at the cells surface, which should affect light intensity less than S1 despite equal ratios. Table 1 lists the extracted parameters at specific time points. The relative change in light intensity per light path length is a constructed parameter that should correlate with the loaded mass (concentration) of the protein in a suspension.

**Table 1:** List of measured and extracted parameters.

| Sample description | Light Intensity [a.u.] | Length of the Light Path [mm] | Mass of Protein Tested [μg] | $\Delta I_r$ per length [%/mm] |
| --- | --- | --- | --- | --- |
| S1B | 21215 | 0.11111 | 1 | 104 |
| S1B + S2F | 21080 | 0.22222 | 1.0001 | 55 |
| S1B + S2E | 21265 | 0.33333 | 1.0011 | 34 |
| S1B + S2D | 21785 | 0.44444 | 1.0111 | 21 |
| S1B + S2C | 23440 | 0.55556 | 1.1111 | 4 |
| S1B + S2B | 23875 | 0.66667 | 2.1111 | 0.8 |

Figure 3(d) shows the change in relative light intensity divided by the light path length versus the total mass of the tested samples. As shown in Table 1, it reveals that as the mass of the protein increased in our experimental system, the intensity of light also increased, revealing that length of the light path was directly proportional to the amount of protein in the sample.

Figure 4(a) and (b) illustrate the definition of the light intensities and light path length. The smart mobile integrated light source emits a light intensity (*I*_*0*_) that is the maximum intensity that can be measured in this experimental setup. The blank intensity (*I*_*b*_) is the measured intensity that goes through the empty container responsible for holding the sample, such as the microcentrifuge tube. The instantaneous measured intensity (*I*) is the recorded light when it passes through the sample. This amount of light intensity strongly depends upon the buffer in which the sample is solubilized/dissolved in, its composition, the light path length, the kind of the suspended analytes and its size in the buffer. The light path length depends upon the loaded amount of suspension inside the container. The path length varies from zero up to the container length (*L*). For a sample with a specific volume (*V*), the corresponding path length is equal to the volume over the cross-sectional area of the container (*A*). Equation (1) expresses the relationship between the relative change in light intensity per light path length and loaded mass (*m*), as follows:

**Figure 4:**
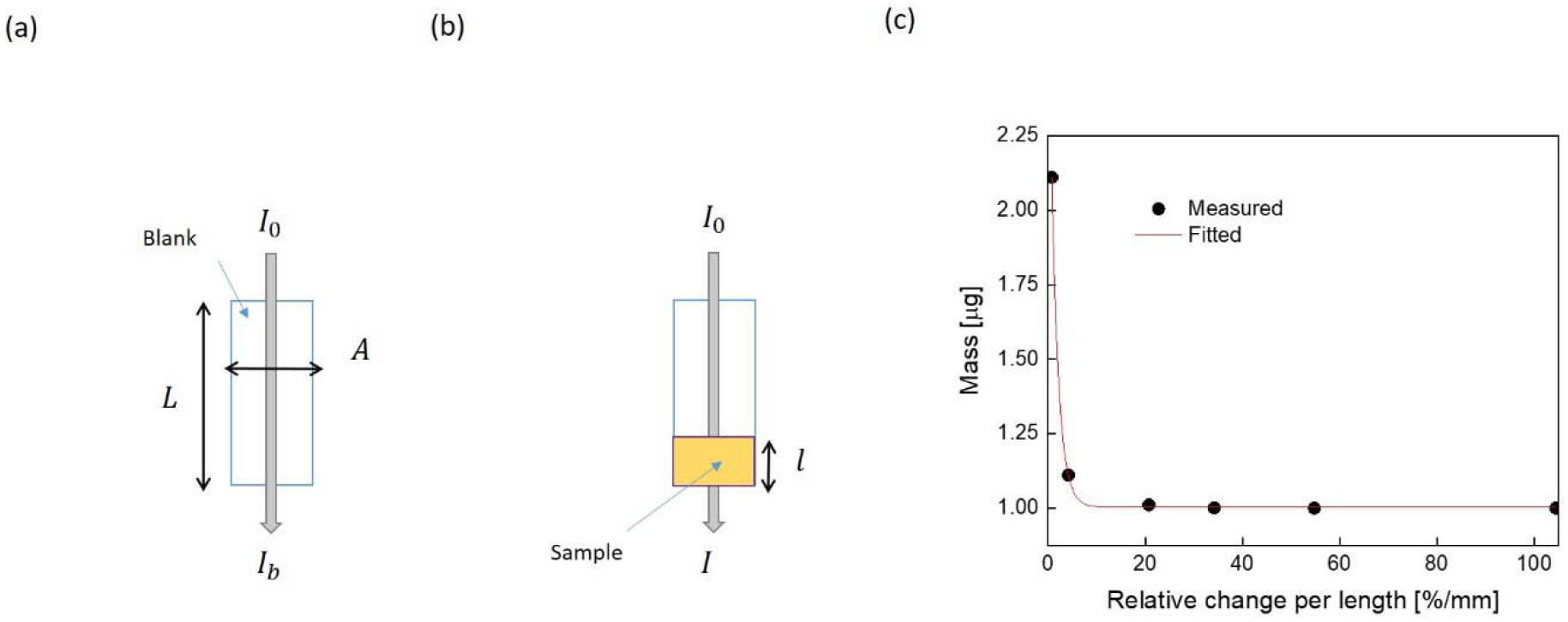
Illustration of light intensity and its path length: **(a)** the blank representation, and **(b)** light path length of the sample. *L* and *A* are the length and cross-sectional area of container, *l* is the light path length. *I*_0_, *I*_*b*_ and *I* are the incident, blank and instantaneous sample intensities, respectively. **(c)** Loaded mass *versus* relative change in light intensity per light-path length. The measured points were fitted with exponential function expressed by equation (1) with the following parameters: m_i_= 1.003μ ± 2.68n, m_f_= 2.163 μ ± 34.7n and α-factor is 1.28435 ± 0.030. The other fitting model accuracy parameters are: Reduced Chi-Sqr, R-Square (COD), Adj. R-Square are 28.8 atto, 1 and 1, respectively, which indicates the best possible fit.

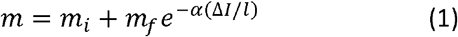

Where: *m*_*i*_, *m*_*f*_ and *l* are the initial mass of the buffer, the mass of the final suspension composite, and the light path length, respectively. α is the decay factor, unique for each control buffer. Its unit is in mm and could be correlated with the material absorptivity. Δ*I* is the relative change in light intensity expressed as follows:

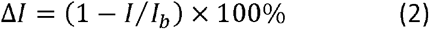

where: *I* and *I*_*b*_ are the instantaneous measured light intensity of the suspension and the corresponding blank, respectively. Figure 4(c) shows the relationship between mass and the relative change per length after fitting the measured points with the exponential function. As can be seen, with more sample volume the path length increases and light intensity decreases; hence, the relative change decreases dramatically.

### Test of Binding Interactions between Spike and ACE2 Using the Optical Assay

After successful demonstration that our set up could detect spike proteins in solution using light, we asked if light intensity could be used to characterize the binding interactions of the spike protein with the viral receptor ACE2. Towards this end, two different variants of the S1 subunit of the spike protein, S1X and S1Y were tested (one form that could bind ACE2 with a much stronger affinity than the other one), along with a non-specific control protein, bovine serum albumin (BSA) that should not bind to ACE2. These proteins were selected to demonstrate the detection of the binding process with ACE2 over time. The measurement process started with the blank, and after 200 seconds, 250 μL of ACE2 protein suspension was tested (Fig. 5(a)). This process was repeated for S1X, S1Y, and BSA and their responses to light were measured individually in the same manner as ACE2. The corresponding individual profiles of ACE2, S1X, S1Y and BSA are depicted in Fig. 5(a) which showed a straight constant line over time. Next, each protein was mixed with the ACE2 separately to detect any possible binding effect. The measurements started with first loading the ACE2 in the blank container, then after 200 ms, the test protein was added to the ACE2 in solution. The responses of the various protein mixtures were read over a period of 15 minutes and are shown in Fig. 5(b).

**Figure 5:**
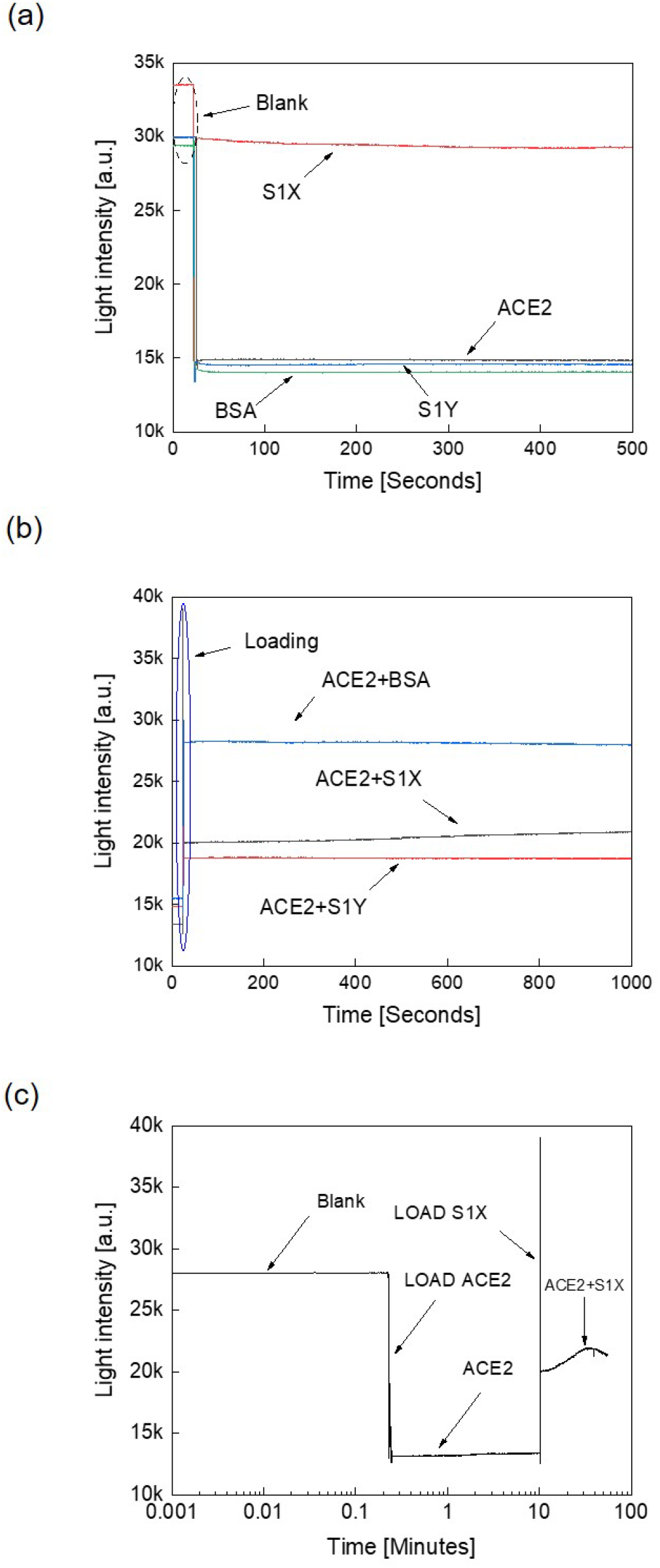
Optical detection of binding interactions between ACE2 and other proteins. **(a)** Measured light intensities over time for individual assessment of ACE2, S1X, S1Y, and BSA. **(b)** The measured mixed light intensities *versus* time for ACE2 mixed with either S1X, or S1Y, or BSA. **(c)** The measured ACE2-S1X interaction profile for an extended time period.

Figure 5(b) shows the corresponding slopes that represent the change of the light intensities over time. The ACE2+BSA and ACE2+S1Y responses exhibited almost constant lines, suggesting highly reduced or lack of any interaction as observed when the proteins were tested individually Figure 5(a). However, the ACE2+S1X profile showed a linear straight line with the maximum-recorded slope. The corresponding light intensity line increased over time, suggesting an interaction between the SIX protein and the ACE2 receptor. We interpret this to mean that there was no protein-protein interaction if the slope of the line was zero; otherwise protein-protein interaction occurred. Based on these observations, our results suggest that the S1X protein exhibits stronger interactions with ACE2, while BSA and S1Y had weaker interactions with ACE2. These observations are confirmed by the fact that whereas S1X has a higher affinity for ACE2 (2 µg/mL S1X can bind 1.5-15 ng/mL ACE2), while S1Y reportedly has a much lower affinity (2 µg/mL S1B binds 0.5-8.7 ng/mL ACE2), as tested in enzyme-linked immunosorbent assays (ELISA) by the company that synthesized these proteins^38^.

To explore the interaction and binding characteristics between ACE2 and S1X in more detail, the measurement time between the two proteins was extended over one hour, the results of which are plotted in Fig. 5(c). As can be seen, a nice “hump” was observed as an increase in arbitrary units (a.u.) with time that was not observed in the other protein mixtures tested which we feel is indicative of the binding reaction between the two proteins.

### Validation of the Optical Assay Using Known Antigen/Antibody Pairs

Next, we wanted to confirm our observations by using our optical system to detect protein-protein interactions using proteins that are well known to interact with each other. This was addressed by testing the molecular interactions between an antigen and an antibody which is similar to the interaction between the spike protein and its receptor. Towards this end, two proteins were tested along with their specific antibodies: the first protein was the receptor binding domain (RBD) of SARS-CoV-2 spike protein and its antibody and the other was the nucleocapsid protein (NCP) of SARS-CoV-2 and its antibody. Similar to the procedure described earlier, the two proteins were tested individually in our optical assay followed by addition of their corresponding antibodies that were mixed and then tested for their interactions.

Figure 6(a) shows the binding between RBD and its antibody. Upon the addition of the antibody, as observed earlier, an “interaction peak” was recorded that is circled in blue color. Similarly, Figure 6(b) shows the binding between NCP and its antibody. However, in this case, we realized that the binding effect occurred at specific antibody concentrations; thus, when the antibody was added first, no interaction peak was observed. Therefore, we added more concentrated antibody and upon its addition, the interaction peak was observed. Addition of more antibody did not allow detection of further interaction peaks, revealing that the protein-protein interaction took place at a specific concentration and once the interaction had taken place, no further interaction took place. For a virus-based suspension, it is therefore suggested to use a fixed antibody concentration and serially dilute the virus suspension to conduct the binding measurements. Certainly, at a specific virus concentration, binding effect will appear in the form of an optical response.

**Figure 6:**
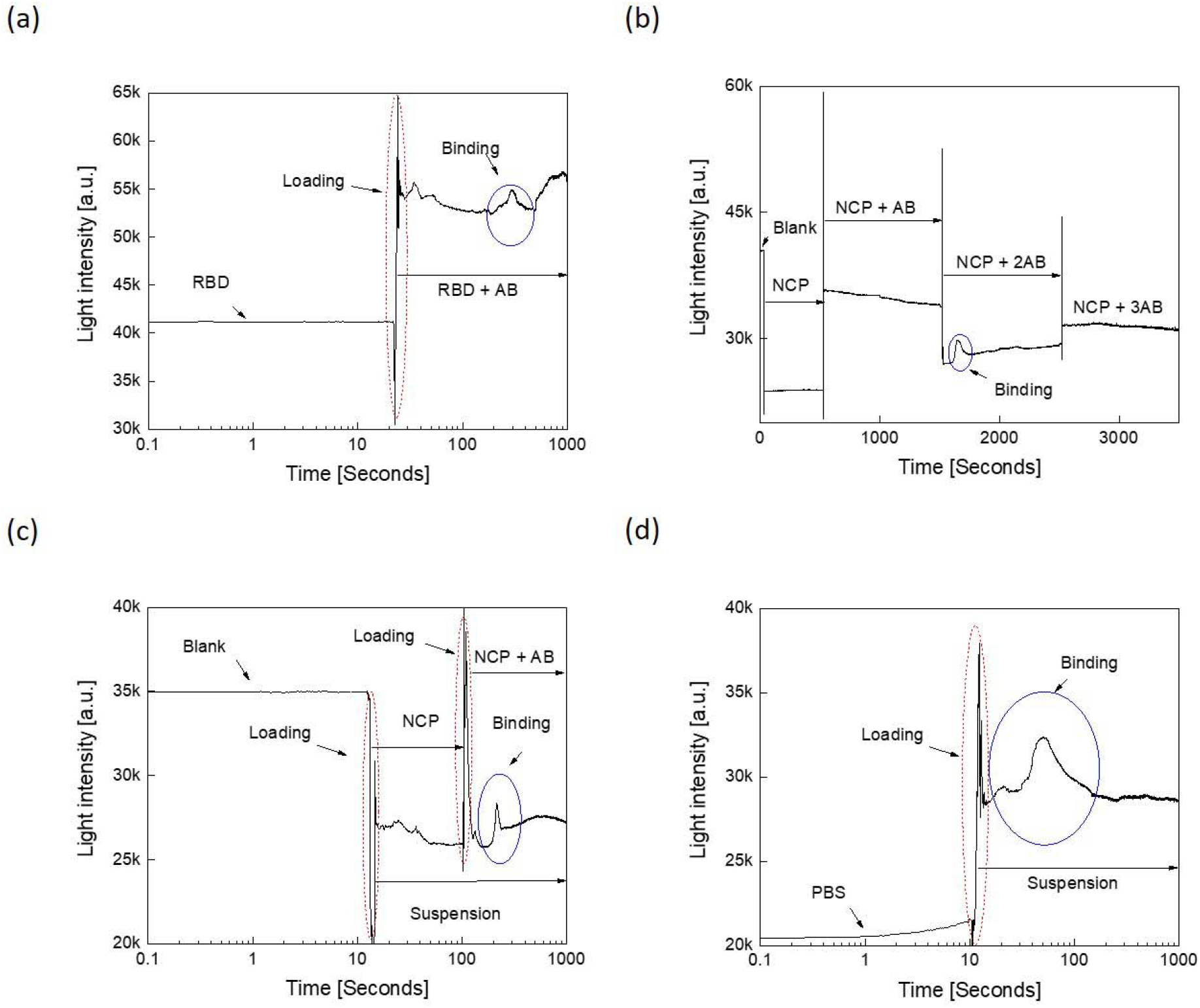
Optical detection of the binding affinities between: **(a)** the receptor binding domain (RBD) of the spike protein with its antibody (AB), and **(b)** the nucleocapsid protein (NCP) and its antibody. The antibody was added again to NCP since no binding interaction was observed the first time. To confirm the result, the antibody was added a third time, but this time once again, the binding interaction was not apparent. **(c)** NCP binding with the antibody after mixing inside, and (d) NCP binding with the antibody after mixing outside. The interaction peak is circled.

Figure 6(c) and (d) illustrate the corresponding optical responses for NC protein and its corresponding antibody when they were mixed either inside (Fig. 6(c)) or outside (Fig. 6(d)) the microcentrifuge tube, respectively. Inside mixing means that the protein was added to the tube and the antibody was added after 10 seconds, while in the outside mixing scenario, both the protein and antibody were mixed prior to being loaded in the tube for optical measurements. As can be seen, the binding response could be detected in each case in the form of appearance of the hump. However, this “hump” was a lot more pronounced when the protein and the antibody were mixed prior to testing than when they were added sequentially. This is good news for the real-life scenario where in a patient sample, the antibody should be already bound to the viral or bacterial antigen at the time of detection.

### Test of the Optical Detection Assay Using a Solid Support

The nitrocellulose membrane is a popular matrix that is frequently used due to its high protein-binding affinity with a pore size of 0.25-0.45 µm in paper-based diagnostics. Protein molecules usually bind to the nitrocellulose membranes through hydrophobic interactions^39^. Due to the ease of their handling, cheap cost, and the presence of hydrophobic interactions between them and the suspended proteins, we tested whether the binding between the SARS-CoV-2 spike protein and antibody could be detected optically when both were added to each other on the nitrocellulose membrane. Using the experimental setup detailed in Fig. 2(a), the optical responses for nitrocellulose membrane, nitrocellulose membrane and spike protein alone, nitrocellulose membrane and antibody against spike protein alone, and nitrocellulose membrane spike protein-antibody were measured. Figure 7(a) shows that both the antibody alone and spike protein alone exhibited higher light intensity than the nitrocellulose membrane alone with almost a straight line with a constant slope over a time period of 10 seconds. The on-paper measured optical responses exhibited fluctuations as in the samples measured using microcentrifuge tubes. This implies that these fluctuations are not due to any interactions; rather, they are due to the spectrometer conversion process^40^.

**Figure 7:**
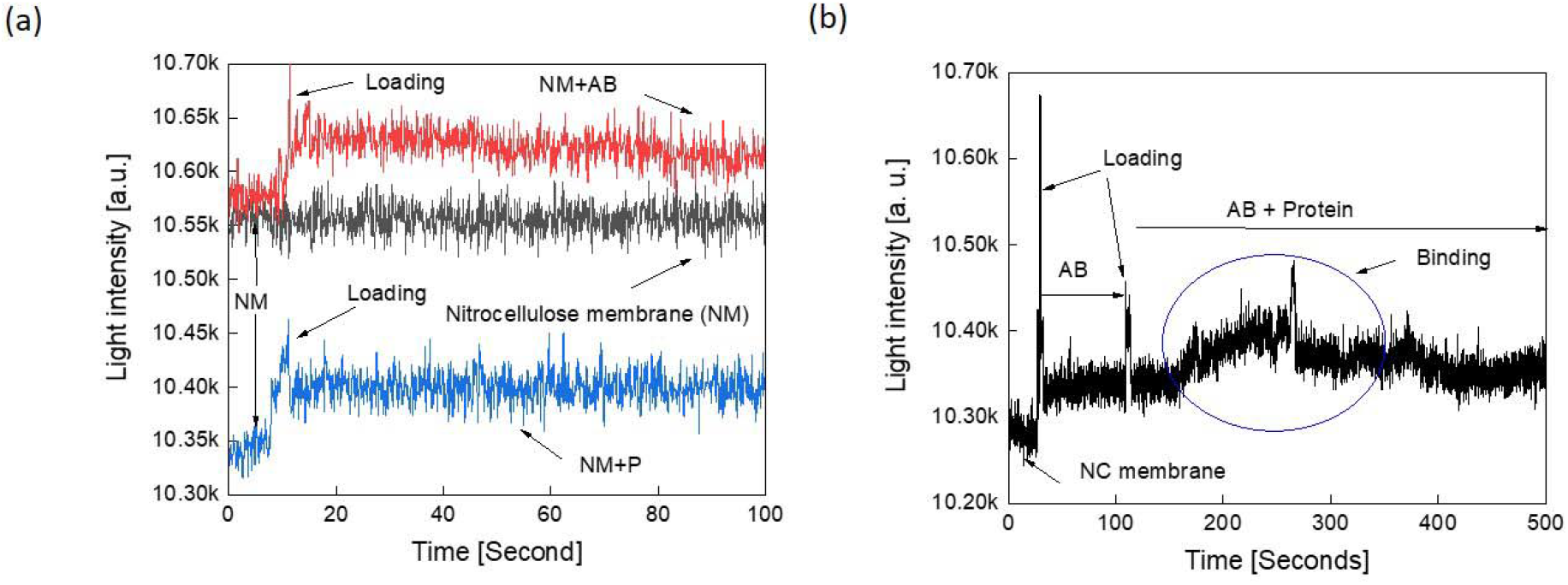
Test of protein-protein interaction measurements on solid support: **(a)** Optical responses on nitrocellulose membrane (NM) alone, nitrocellulose membrane and spike protein (NM+P), and nitrocellulose membrane and antibody to spike protein (NM+AB) alone. **(b)** Optical responses to spike protein-antibody binding on the nitrocellulose membrane.

Figure 7(b) summaries the interaction measurements which start with the membrane alone. After 20 seconds, the antibody suspension was loaded on the membrane and measurements were conducted up to 100 seconds. Next, the spike protein sample was loaded and measurements were continued up to 500 seconds. As revealed from Fig. 7(b), the interaction peak clearly appeared as circled in blue. It is worth noting that the membrane size, shape, and charge of biomolecules, pH and viscosity of the control buffer, as well as the composition influences the corresponding optical response and binding interactions and must be carefully standardized^41^.

### Role of Electric Current in Disrupting Protein-Protein Interactions

Finally, we studied the effect of direct current (DC biasing) on the ability of two proteins to bind specifically. This was achieved by subjecting the NC protein solution to DC voltage bias, as depicted in Fig. 8(a). An applied bias should result in an induction of current across the suspension. If this current is high enough, it should have the potential to destroy the protein physiology and functionality, resulting in the loss of specific protein-protein interactions. To test this hypothesis, the NC protein solution was loaded in an electroporation cuvette (rather than a microcentrifuge tube) that incorporates two electrodes with a volume of 0.5 mL and a separation distance of 0.4 cm. This should result in a breakdown electric field of 7.5 V/cm. At this field onwards, the binding between the protein and the antibody should be affected. Above this field, the sample should be incapacitated for binding. Figure 8(a) reveals that the optical response decays slowly with the application of DC bias. At 3 volts DC bias, the optical response decays with a considerable step, and increasing the DC bias further should burn the suspension and destroy it.

**Figure 8:**
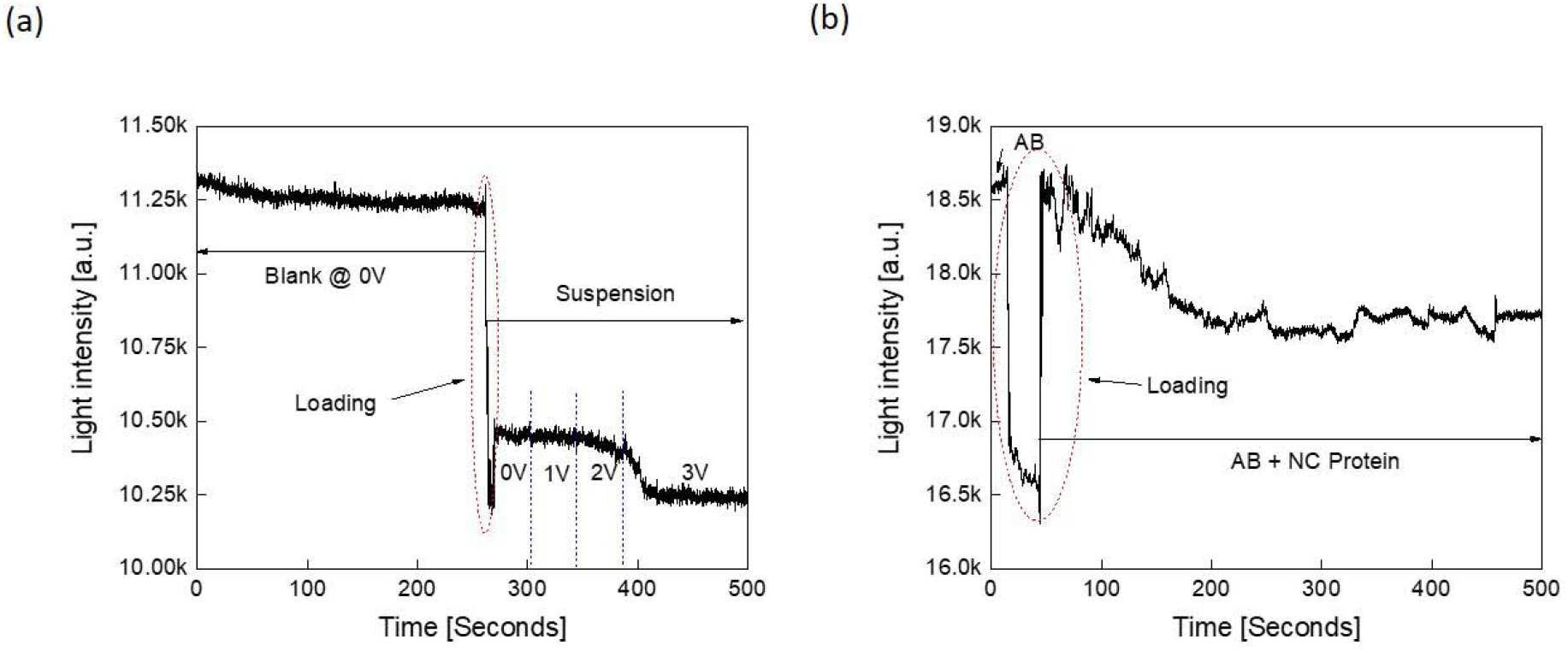
Opto-electrical measurements: **(a)** Measured NC protein optical response *versus* time at different DC bias voltages. **(b)** Binding measurements between NC protein and its corresponding antibody after subjected the solution to an electric field.

The breakdown field depends on the electrical characteristics of both the buffer and the analyte such as proteins, viruses, etc. To explore this further, the suspension of protein was subjected to 3 V for 1 minute and then the antibody to NC was added to the NC protein solution. The corresponding measured response is shown in Fig. 8(b). The measured response was observed to be noisy and did not show a clear binding effect when compared with Fig. 6(c) that reported the optical response for the same protein and antibody without the application of DC bias. It is worth mentioning that it may be possible to create a corresponding vaccine for a disease by subjecting the target viral protein to DC bias which will affect its function and destroy its physiology (denature it) and communicability (binding interactions). Furthermore, the proposed optical detection in time domain can also be used for monitoring and detecting the efficiency of vaccine process development.

## Discussion

This study establishes the proof-of-principle that optical methods can be used to detect specific SARS-CoV-2 spike proteins or their subunits (S, S1, & S2) as well as their interactions with the ACE2 receptor in solution, whenever present (Figs. 3 & 5). The principle was further validated by testing specific protein-protein interactions by testing two viral protein-antibody pairs (RBD and NC proteins with their specific antibodies) and testing them either in solution (Figs. 6) or on a solid matrix (Fig. 7). Finally, it was shown that application of a weak current into the system could lead to the disruption of NC protein-antibody interactions which could be optically detected (Fig. 8). In other words, our technique could be used not only to detect specific SARS-CoV-2 spike protein-receptor interactions, but also result in destruction of protein interactions important for virus replication; thus, inhibiting rate of infection. The proposed detection method can be performed within minutes, without the need to biochemically label the proteins. This system can be used to develop novel optical-based detection tests for any virus in a specific and sensitive manner as long as one specific protein partner is available in solution or on a solid support that can interact specifically with a specific viral protein. For instance, in the case of SARS-CoV-2, one could use either an antibody to the spike protein or the ACE2 protein to determine whether a particular patient sample may have the virus.

Figure 1(c) shows the spikes distribution suspended in a sample. As illustrated, the spike proteins are randomly distributed and exhibit Brownian motion by default^42^. The same scenario applies to the distribution of ACE2 as illustrated in Fig. 1(d)^43^. If specific binding occurs between the spike proteins and ACE2, as represented by Fig. 1(e), their binding distribution should still exhibit random Brownian motion. Nevertheless, SARS-CoV-2 spike protein binds with human ACE2 protein with a specific binding energy that has been measured and is estimated to be nearly −58.55 ± 8.75 kcalmol^−1 44,45^. A conformational change occurs in ACE2 receptor protein after binding with spike protein fragment^46^. Ov et al. have shown that the refraction index changes due to the binding interactions after virus-antibody incubation process^47^. To detect binding of living cells and viruses with potential drugs, they have proposed a novel label-free real time approach incorporating a long range surface waves on one-dimensional photonic crystal surface along with microfluidic channel technology^47^.

The photoelectric effect theory combines kinetic energy, binding energy and photon energy all three of which are correlated through the theoretical physics fundamentals and principles^48^. Hence, if the generated photon energy due to the binding interactions is sufficient, it could give rise to light intensity at a specific wavelength^49^. Wang et al. have discussed the enhancement of receptor binding of SARS-CoV-2 through networks of hydrogen-bonding and hydrophobic interactions^44^. They have provided explanations to better understand the structural and energetic details responsible for protein–protein interactions between the host receptor ACE2 and SARS-CoV-2. Their simulations reveal that both electrostatic complementarity and hydrophobic interactions are critical to enhance receptor binding and escape antibody recognition by the RBD of SARS-CoV-2. Ortega et al. have conducted an in silico analysis to study the role of changes in SARS-CoV-2 spike protein during interaction with the ACE2 receptor. They have concluded that the binding energy generated during the SARS-CoV-2 spike and ACE2 interactions can be reduced due to mutations in the sequence of the spike protein^18^. Dahal et al. have demonstrated that binding probability increases with antibody concentration and the stability of protein^50^.

Based on these observations, we believe that a “hump or spike” in light intensity is observed when a specific molecular interaction takes place between two proteins. This is mainly due to the physio-chemical properties of the proteins that relates to binding affinity in the contact surface area which incorporates the association/dissociation process^51^.

As revealed from the corresponding binding measured light intensity profiles, they exhibit Gaussian-like peaks. Wang et al. have demonstrated in their study that the molecular binding at the single molecule level displays such a peak^52^. Interestingly, Kozono et al. monitored the real-time Brownian motion and fitted it with Gaussian function^53^. The fitting parameters of the distributions can provide many features of the binding interactions^54^. This could provide a quantitative signature or characterization of a specific antigen binding to a specific antibody such as intrinsic specificity and binding rate. It is also proven that the probability of the binding free energy to be Gaussian distributed near the mean and exponential-like distributed in the tail^55^. Figure 9 shows the binding interaction over the corresponding time intervals for RBD and NCP proteins with their corresponding antibodies under different conditions as described in Fig 6. Similar interactions peaks were observed in each case except they varied in the time of appearance and extent of light intensity. The time slot denoted by (i) represents the time just before the interaction occurs. As the interaction starts, the corresponding optical profile ascends incrementally as indicated by (ii) due to the increase in binding events, releasing more photons energy. The peak pointed by (iii) occurs at the maximum event of binding between antigens and their antibodies. The height of the peaks indicates stronger interactions and *vice versa*. The profile then descends till the end as the binding events becomes less and no further interactions occur at the end, as illustrated by (iv). The distance in time to maximum peak reflects the speed of the binding interactions; thus, the earlier the peak appears, the faster the binding interaction takes place. As can be seen in Fig. 9, the speed of NC protein binding to its antibody took place between 100 and 1000 seconds irrespective of the dilution of the proteins.

**Figure 9:**
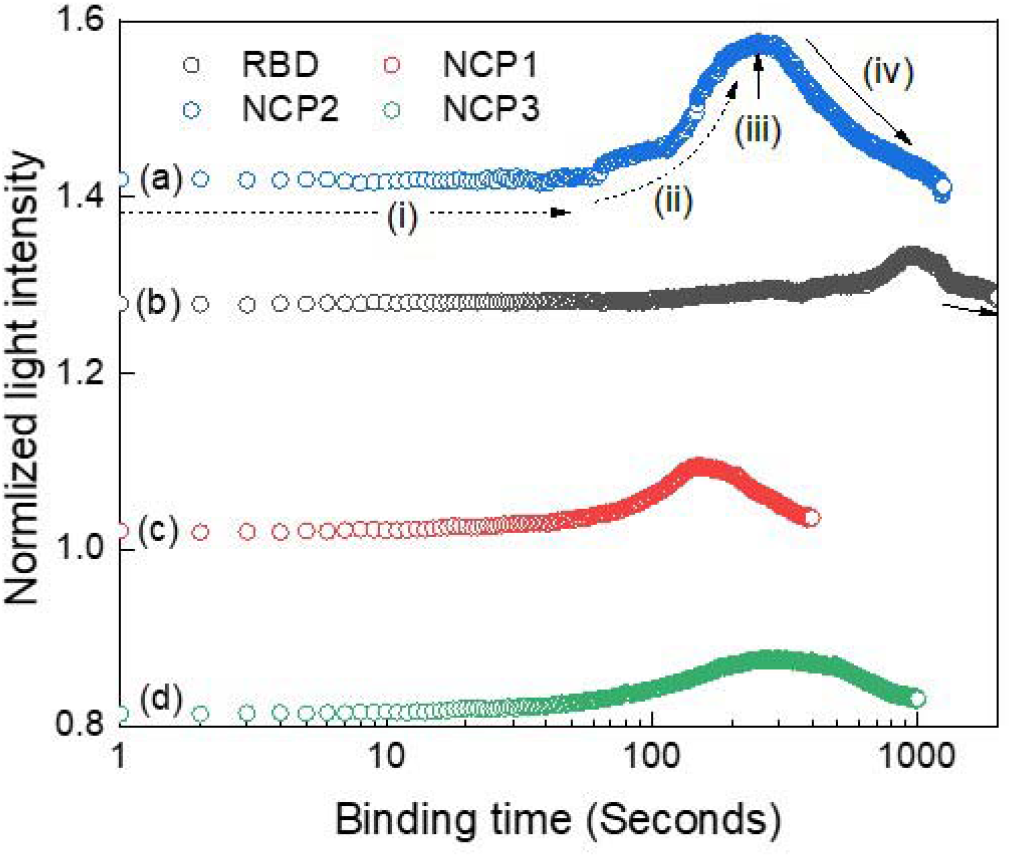
Optical profiles for normalized interactions *versus* binding time under different conditions for RBD and NCP. (a) NCP with its antibody at the maximum concentration of 22 μg per ml (b) RBD with its antibody, (c) NCP with its antibody at a 1:10 dilution (d) NCP with its antibody at 1:10 with outside mixing.

Next, we analyzed the interaction profiles of these samples further by fitting them to Gaussian function. Table 2 lists their corresponding fitting parameters. The most important parameters are the width and the maximum peak amplitude. Base and center parameters represent the offset level and the maximum peak location, respectively. These two parameters provide minor information and could be set to fixed values such as zero in all profiles. The multiplication of the maximum amplitude with its corresponded width can be utilized as an indicator to describe the speed of interaction. This “indicator” has been listed in Table 2, last row. Accordingly,NCP2 exhibited the fastest binding, while NCP3 exhibited the slowest binding with the arrangement from fast to slow being as follows: NCP2, NCP1, RBD, and NCP3.

**Table 2:**
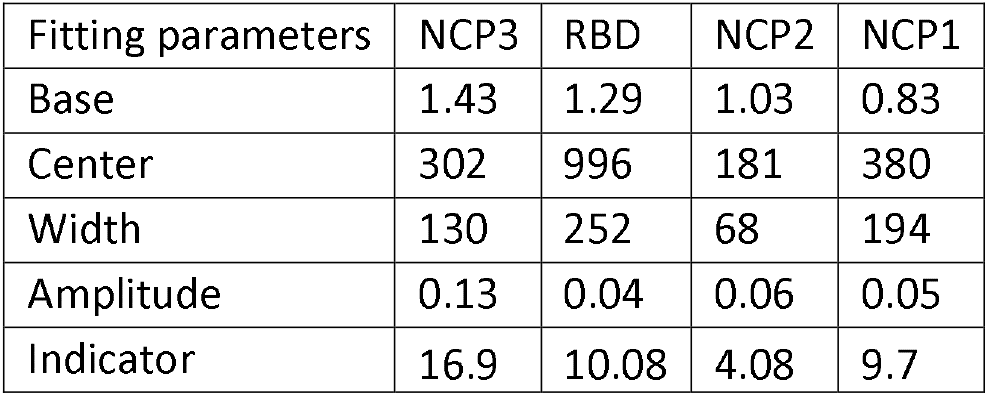
Fitting parameters for interaction profiles depicted in Fig. 9.

### Practical Applications of the Proposed Technique

The optical approach presented in this study can be easily turned into a functional working system to detect SARS-CoV-2 or any other viral or bacterial pathogen against which an antibody is available that can detect it sensitively. Currently, there are several available handheld portable spectrophotometers which are compact and lightweight, and equipped with wireless communication system for data transmission. The data can be received by the smart phone through Bluetooth or Wi-Fi technologies equipped with a mobile app designed to process the received optical profiles over time. The collected optical profiles *versus* time can then be processed immediately using the smart phone processor and computational resources. The results can be displayed on the same smart phone immediately as well. The antibody could be coated on flexible strips and kept inside packs with medium. These strips can be used directly for loading the nasal swab in-position within a fabricated holder using 3D printing technology. The 3D printed holder can be designed to integrate the portable sensor and the smart phone as well. The concept of strips can be further used to detect different specimens taken from blood, breath, urine, nasal swabs, stool, etc. For example, the subject can breathe, exhale, or sniff into a device with a probe coated with antibodies. If binding occurs, the integrated device should be able to pick up the interaction, confirming that the patient is infected with the virus to which the antibodies are directed against.

The current methodology provides a rapid, reproducible and accurate detection mechanism that can be used to create home-based COVID-19 detection tests that can be used by anyone. The authors envision that such tests would not require any laboratory setting, and could be performed using test strips coated with antibodies without prior sample processing. Interestingly, our approach does not require any electrode patterning which makes it the best fit for massive production and high-volume use. It can be adapted as a point-of-care testing platform with high-throughput to be used in schools, airports, malls and public services places as well. The cost per test is expected to be less than one dollar which will make it competitive with current market price. The detection platform can be easily equipped with standard electronic data transmission systems to transmit and process data in place and share it with family, doctors and hospitals over wireless transmission. Furthermore, the platform can be deployed in high-risk areas with ease-of-use and clear steps to load the specimen and easy to understand instructions to operate.

Furthermore, when compared with other detection concepts and methodologies, the presented approach can distinguish between influenza and coronaviruses. ACE2 binds directly to the viral spike protein, while ACE2 plays an important role in acute lung injury induced by influenza viruses^56^ which can be correlated with disease severity^57^. Hence, the proposed methodology can be used in reverse where the spike protein can be used for ACE2 detection.

## Conclusions

In summary, this study provides the proof-of-principle for an optical-based, quick, simple, and sensitive screening technology for the detection of SARS-CoV-2. It is based on the principle that when light passes through a sample, interactions between the photons and sample occurs within a specific range of frequencies. The current approach utilizes a smartphone light source and a portable mini-spectrophotometer to convert the variations in light intensity into measured signal. The optical detection exhibited high sensitivity towards selected SARS-CoV-2 proteins with a low detection limit which was five times more sensitive than published real time quantitative PCR assays. The optical responses could further be analyzed using the principle component analysis technique to enhance and allows precise detection of the specific target in a multi-protein mixture. This approach can be further developed to accommodate mass screening that should provide fast and accurate positive or negative test results.

## Data Availability

NA

## Acknowledgements

We thank the Research office at UAEU for their continuous help and support.

## Author contributions

MA conceived the study. MA, FM and TAR designed the experiments. MA and NP performed the experiments. MA, FM, and TAR analyzed the data. All authors reviewed and edited the paper.

## Materials and methods

### Optical Mini-Spectrometers

C11708MA from Hamamatsu/Japan^36^ was used to convert the variable attenuation of light waves as they passed from end-to-end or reflect off substances into signals with spectral response ranging from 640 to 1010 nm. The wavelength reproducibility ranged between −0.5 to 0.5 nm and had a maximum of 20 nm FWHM spectra, under constant light conditions. The measurements were conducted with the room lights on. The distances between the light source, the spectrometer, and the sample holder were adjusted to eliminate any possible interference and to stabilize the spectrometer performance. Furthermore, the spectrometer was aligned with the light source and sample cuvette to achieve a straight path of light.

### Smart mobile phone

The smartphone light source was used as the main light source. The mobile light emits lights with the spectral range from 380 to 740 nm. The maximum optical power was emitted at a wavelength of 623 nm. In this work iPhone 8 was employed, though any smart phone can be used.

### Electroporation Cuvette

The 0.4 cm-gap MicroPulser Electroporation Cuvettes from Bio-Rad were used for the electrical analysis^58^. This a high-quality cuvette which is compatible with most electroporation systems. The cuvette incorporates aluminum electrodes plates with an area of 1 cm by 0.8 cm. Its outer dimensions are 12.5 x 12.5 x 45 mm (W x D x H) with a path length of 10 mm and a functional volume between 50–1,500 µl.

### Gamry Reference 3000™

It is a high-performance potentiostat/galvanostat/ZRA that was used to apply the constant DC biasing voltage required in this work^59^. The Gamry analyzer exhibits a high-current, high-performance potentiostat, fully equipped to perform impedance spectroscopy up to 1 MHz. The analyzer is able to measure current with an accuracy of ±5 pA with voltage applied accuracy of ±1 mV. Gamry 3000 instrument is equipped with a wide range of electrical measurement capabilities. The analyzer should be calibrated using the manufacture-provided calibration kit to exclude the effect of lengthy cables.

### Nitrocellulose membranes

Nitrocellulose is a popular matrix used due to its high protein-binding affinity with a pore size of 0.25-0.45 µm in paper-based diagnostics^60^. Protein molecules bind to the nitrocellulose membranes through the hydrophobic interactions. The nitrocellulose membranes are easy to handle and are cheap. The membrane size, shape, and charge of biomolecules, pH, viscosity and ionic strength of the control buffer, as well as the composition influences the electrical response. Excessive electrical field may cause a solution to deteriorate and stick to the membrane and cause joule heating which will decrease the resistance of the control buffer and lose its buffering capacity and deter the electrical performance of the capacitance structure.

### Spikes proteins

The SARS-CoV-2 S1 (ProSci, Cat.No.97-087)^38^ and S2 (ProSci Cat.No.97-079)^38^ recombinant proteins were purchased in lyophilized form and resuspended according to the manufacturer’s instructions at a stock concentration of 600 µg/ml for S1 and 200 µg/ml for S2 protein in sterile water. Two different spike S1 recombinant proteins (S1X (ProSci Cat no. 10-109) and S1Y (ProSci Cat no. 10-111)) ^38^were also purchased in the lyophilized form. The binding affinity of these two S1 proteins varies due to the difference in the amino acid sequence that complements specific binding to the ACE2 receptor.

### ACE2 receptors

The human ACE2 recombinant protein (ProSci Cat No. 10-114)^38^ was resuspended in sterile deionized water at a stock concentration of 500 ng/µl as per the manufacturers instruction and used for the analysis.

### Nucleocapsid protein

The SARS-CoV-2 nucleocapsid protein (Sino Biologicals, Cat no. 40588-V08B)^61^ and its corresponding nucleocapsid antibody (Sino Biologicals Cat no. 40588-T62)^61^ were used for the binding affinity experiments. The lyophilized protein was resuspended at a stock concentration of 0.25 mg/ml according to the manufacturer’s instruction in sterile water. The nucleocapsid rabbit polyclonal antibody was supplied at a stock concentration of 1 mg/ml.

### The receptor binding domain (RBD)

The receptor binding domain (RBD) of the SARS-CoV-2 spike protein (Sino Biologicals, Cat No 40592-V05H)^61^ was expressed as a recombinant protein with the Fc region of mouse (mFc) at the C terminus end and its corresponding spike RBD antibody (Sino Biologicals, Cat No 40592-T62)^61^ was used for the binding affinity experiments. The RBD protein was prepared in sterile water at a stock concentration of 0.25 mg/ml, as per the manufacturer’s instruction. The spike RBD rabbit polyclonal antibody was prepared at a stock concentration of 1 mg/ml and diluted further for analysis.

